# Frequent suboptimal thermocycler ramp rate usage negatively impacts MTBDR*sl* performance for second-line drug resistant tuberculosis diagnosis

**DOI:** 10.1101/2021.05.18.21257375

**Authors:** Brigitta Derendinger, Margaretha de Vos, Samantha Pillay, Rouxjeane Venter, John Metcalfe, Yonas Ghebrekristos, Stephanie Minnies, Tania Dolby, Natalie Beylis, Robin Warren, Grant Theron

**Affiliations:** DSI-NRF Centre of Excellence for Biomedical Tuberculosis Research, SA-MRC Centre for Tuberculosis Research, Division of Molecular Biology and Human Genetics, Faculty of Medicine and Health Sciences, Stellenbosch University, South Africa; Division of Pulmonary and Critical Care Medicine, Zuckerberg San Francisco General Hospital, University of California San Francisco, USA; National Health Laboratory Service, Groote Schuur Hospital, Cape Town, South Africa; National Health Laboratory Services, Green Point, Cape Town, South Africa

## Abstract

Strengthening the detection of second-line drug-resistance is a key tuberculosis (TB) control priority. The performance of MTBDR*plus*, a multidrug-resistant (MDR)-TB assay is reduced when suboptimal ramp rates are used. We investigated ramp rate’s effect on MTBDR*sl*; the most widely-used molecular second-line drug-resistant TB assay.

We tested 52 smear-negative Xpert MTB/RIF Ultra-positive sputa and a *Mycobacterium tuberculosis* (*Mtb*) dilution series at manufacturer-recommended (2.2°C/s) or most common suboptimal ramp rate (4.0°C/s; identified via an earlier survey). *Mtb-*complex DNA (TUB-band)-positivity, indeterminate rates, fluoroquinolone- and second-line injectable-resistance accuracy, banding differences and, separately, inter-reader variability were assessed.

39% of re-surveyed laboratories (5/13) did not use the manufacturer-recommended MTBDR*sl* ramp rate. On sputum, this ramp rate improved indeterminates vs. 4.0°C/s (0/52 vs. 7/51; p=0.006), false drug-resistance calls (0/104 vs. 6/102; p=0.013), and incorrect banding calls (0/1300 vs. 55/1275; p<0.001). Valid results (neither TUB-negative, indeterminate, nor any false drug-resistance calls) (52/52 vs. 41/51; p=0.001) on sputa hence improved by +21% (95% CI: 8-34%) with optimal ramp rate usage. Suboptimal ramp rate increased banding call inter-reader variability [52/1300 (4%) vs. 34/1300 (3%); p=0.030] on sputa but not dilution series; highlighting the importance of using clinical specimens for assay performance evaluations.

Suboptimal ramp rate contributes to poor MTBDR*sl* performance. Ramp rate correction will improve second-line drug-resistant TB diagnoses. Laboratories must ensure the optimal manufacturer-recommended ramp rate is used.

## Introduction

In 2019, ∼10 million people fell ill with tuberculosis (TB) and ∼1.3 million people died^1^. Drug resistant (DR)-TB is a global health problem. ∼465 000 people having multidrug resistant (MDR)-TB, ≥6% of which have additional resistance to (FQs) and second line injectables (SLIDs)^1^. Worldwide only 52% of MDR-TB patients were tested for resistance to both these drug classes and only 58% of those who start treatment successfully complete it^1^. Phenotypic culture-based drug susceptibility testing is slow and costly, and patients can wait six months before placed on effective treatment, if at all^2^.

GenoType MTBDR*sl* (MTBDR*sl*; Hain Lifescience, Germany)^3^ is one of two commercially-available rapid molecular World Health Organization (WHO)-endorsed assays for the detection of *Mycobacterium tuberculosis* (*Mtb*) complex and resistance to FQs and SLIDs^4^. Per the WHO, MTBDR*sl* should be done directly on sputum irrespective of smear microscopy status to reduce the delay associated with culture for indirect testing^4^.

However, performance data for direct use on sputum are heterogenous. In a systematic review and meta-analysis, smear-negative sensitivity estimates were imprecise: 80% [95% confidence interval (CI) 28-99%], 80% (28-99%) and 50% (1-99%) for FQs, SLIDs and XDR-TB, respectively^5^. This affected the certainty of evidence of the WHO recommendation and undermined MTBDR*sl*’s uptake.

MTBDR*sl* requires thermocycling for DNA amplification. The manufacturer recommends a ramp rate of ≤2.2°C/s, which is the speed of temperature change between PCR cycles. We previously showed Genotype MTBDR*plus* (MTBDR*plus*; Hain Lifescience, Germany) performance, which is an assay for first-line resistance, is reduced when suboptimal thermocycler ramp rates are used, most so on smear-negative specimens^6^. These findings are incorporated into laboratory external quality assessment programmes and WHO TB laboratory training material^7^.

If MTBDR*sl* is also vulnerable to this phenomenon, this would result in some of the thousands of people who receive this assay each day having drug resistance diagnoses missed; thereby resulting in resistance to the drugs critical to protect new regimens (e.g., FQ to limit bedaquiline resistance acquisition in the oral second-line regimen) remaining delayed or undiagnosed^8,9^. More broadly, this issue of ramp rate is increasingly pertinent as manufacturers are designing instruments with faster thermocycling (and hence faster ramp rates) to decrease time-to-result. Furthermore, many thermocyclers, especially those at entry-level, do not have a customisable ramp rate.

We hypothesised that the heterogenous and suboptimal sensitivities reported for MTBDR*sl* on smear-negative specimens were partly attributable to suboptimal ramp rate and sought to generate empirical evidence of this. We assessed whether laboratories that reported use of suboptimal ramp rates during our MTBDR*plus* evaluation^6^ had switched to the manufacturer-recommended ramp rate, and what the observed effect had been.

## Methods

### Ethics statement

This study was approved by the Health Research Ethics Committee of Stellenbosch University (N16/04/045) and Western Cape Research Ethics Committee (WC_2016RP18_637). All methods were in accordance with relevant guidelines and regulations. Permission was granted to access anonymised residual specimens collected as part of routine diagnostic practices and thus patient informed consent was waived.

### Experimental design

Ramp rate assessment was done in both an *in vitro* dilution series and clinical sputa **(Figure 1)**. DNA extracted from dilution series and clinical specimens were split and compared head-to-head at the manufacturer-recommended ramp rate of 2.2°C/s or the most common suboptimal ramp rate of 4.0°C/s identified previously in a survey^6^. MTBDR*sl* was done on all amplified DNA per manufacturer’s instructions for use^3^. For sputa, programmatic MTBDR*sl* results (done at the recommended ramp rate) were compared. All equipment is annually calibrated and serviced.

**Figure 1.**
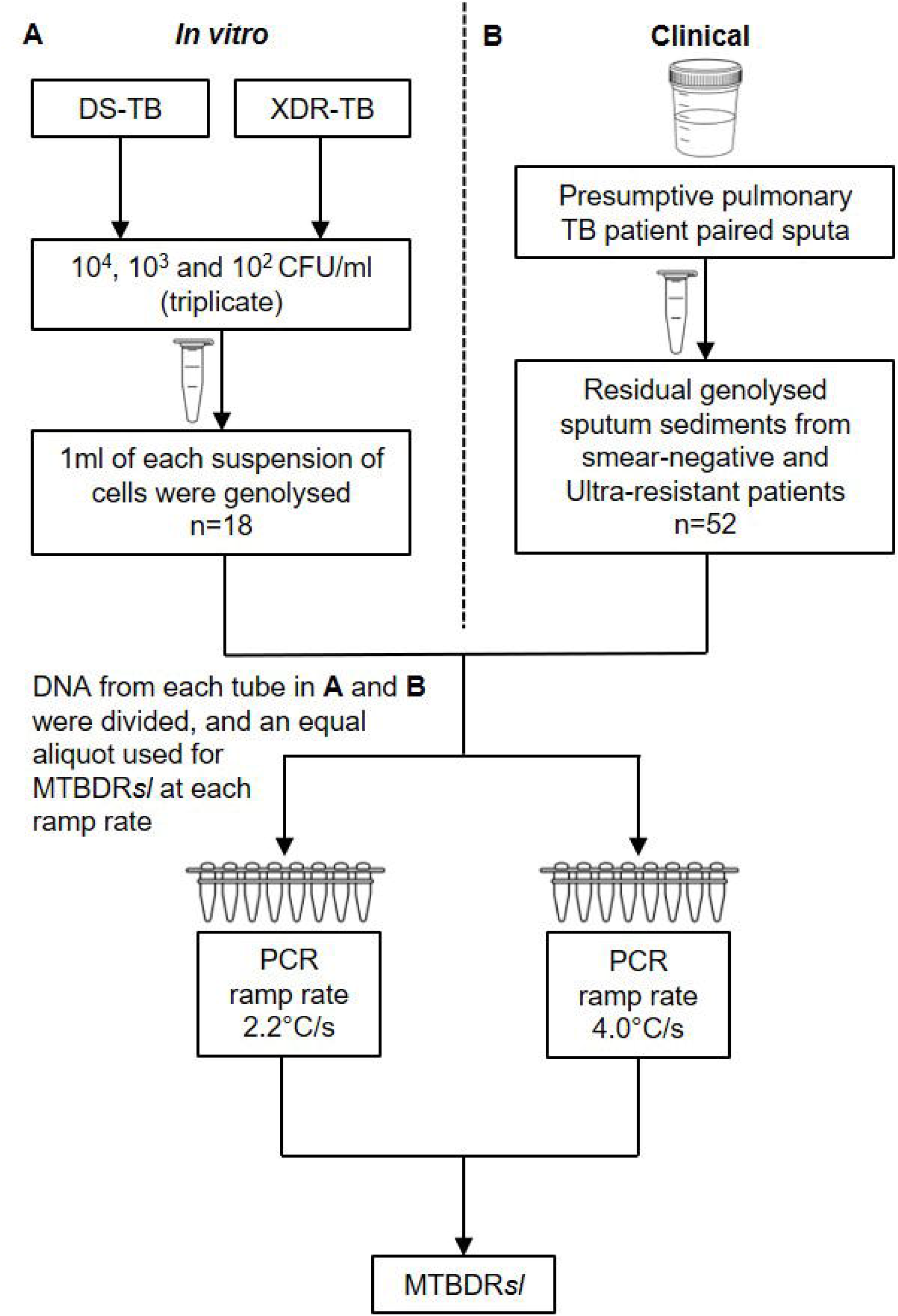
Study flow diagram for (**A**) *in vitro* [a dilution series of cells (10^4^, 10^3^, 10^3^ CFU/ml)] experiment and (B) clinical experiment (sputa) to assess the impact of thermocycler ramp rate on MTBDR*sl*. DNA extracted from the dilution series and clinical specimens was split and MTBDR*sl* compared head-to-head at the manufacturer-recommend ramp rate of 2.2°C or 4.0°C/s. TB □ tuberculosis; DS-TB □ drug susceptible tuberculosis; XDR-TB □ extensively drug-resistant tuberculosis; CFU □ colony forming units; ml □ millilitre

### MTBDRsl calls and result definitions

#### CC band

This must be present for a strip to be valid as it indicates hybridisation occurred.

#### AC band

Present when the assay is done correctly. Per the manual^10^, there are rare cases where the AC band disappears due to competition during the amplification reaction. In this scenario, an absent AC band in combination with TUB and locus control bands is still a valid result.

#### Locus control bands (gyrA, gyrB, rrs, eis)

Need to be present for a call from that locus to not be indeterminate.

#### Positive for Mtb-complex DNA

TUB-band present.

#### Strip banding call

For a band to be classified as present, it must be equal or darker than the amplification control (AC) band. Overall, there are 27 possible strip bands on MTBDR*sl*. When only the CC- and AC-bands are present, this represents a valid TUB-negative result.

#### Diagnostic call

Band presence or absence in a region determines whether the result is classified as susceptible or resistance to a drug class (two diagnostic calls possible for MTBDR*sl*: FQs or SLIDs).

#### (In)determinate for a gene region and/or drug class

For a specific gene region and/or drug class to be determinate, locus control band(s) must be present. We called a strip indeterminate for a drug class if at least one gene locus control was absent.

#### Valid result

TUB-band positive strip determinate for all gene locus controls and thus has diagnostic calls for both drug classes (e.g., TUB-band positive, FQ-resistant, SLIDs-susceptible).

### *Impact of thermocycler ramp rate on MTBDR*sl *performance on a dilution series*

A phenotypically- and genotypically-confirmed clinical XDR strain (known *gyrA, gyrB, rrs* and *eis* variants) and a drug-susceptible (DS) strain (H37Rv, ATCC 25618) were grown to mid-exponential phase (approximately 10^8^CFU/ml) in Middlebrook 7H9 media (Becton Dickinson, United States) supplemented with Middlebrook Oleic Albumin Dextrose Catalase (Becton Dickinson, United States). Serial dilutions in phosphate buffer supplemented with 0.025% Tween 80 (Merck, South Africa) were inoculated onto Middlebrook 7H10 solid media (Becton Dickinson, United States) and incubated for 21 days at 37°C for colony forming unit (CFU) calculations. This was done in biological triplicate. 1ml of the 10^4^, 10^3^ and 10^2^CFU/ml suspensions were GenoLysed (Hain LifeScience, Germany) and MTBDRsl done per the manufacturer’s instructions^3^. The two lower dilutions approximate to smear-negative disease (<10 000CFU/ml)^11^; expected to be most affected by suboptimal ramp rate. DNA was amplified with the CFX96 thermocycler (Bio Rad Laboratories, South Africa) at ramp rates of 2.2°C/s and 4.0°C/s. Two experienced readers recorded bands in a blinded manner. Accuracy analyses for TUB-band positivity, indeterminate rates, incorrect banding calls, and incorrect diagnostic calls were done.

### *Impact of thermocycler ramp rate on MTBDR*sl *on clinical specimens*

Genolysed samples (n=52) remaining after programmatic LPA testing were collected from a TB laboratory in Cape Town, South Africa. These samples were, per the national algorithm, derived from the paired sputum specimen of a presumptive pulmonary TB patient that received Ultra (on a separate sputum), MGIT 960 culture and Auramine O microscopy (on the same sputum later GenoLysed). All sputa were smear-negative and Ultra-rifampicin resistant. We did not include smear-positives as we previously showed ramp rate to not affect MTBDR*plus* on smear-positives^6^. Residual GenoLysed samples were stored at −20°C.

Samples were categorised using programmatic LPA results as: 17 MDR-TB, 24 pre-XDR and 11 XDR-TB. For the experiment, DNA was amplified using a CFX96 thermocycler (Bio-Rad, United States) at 2.2°C/s (manufacturer-recommended) and 4.0°C/s. MTBDRsl was done per the manufacturer’s instructions^3^, and two experienced readers recorded bands in a blinded fashion. Accuracy analyses for TUB-band positivity, indeterminate rates, incorrect banding calls, and incorrect diagnostic calls were done.

### *Calculation of laboratory savings from an improvement in MTBDR*sl *performance om smear-negatives stemming from ramp rate*

We calculated how much the routine laboratory, from which we received GenoLysed remnants, would save if we applied the proportional increase we found in valid results when the optimal vs. the suboptimal ramp rate was used. This cost savings calculation was based on the average number of MTBDR*sl* tests done indirectly on cultured isolates per month (which would now be reduced due to direct testing on smear-negatives having improved performance) and the cost of each test (including consumables, labour, and overheads; the sum is pre-calculated supplied by the laboratory provider).

### Inter-reader agreement

Three experienced readers read all strips from the dilution series and clinical specimens at either ramp rate independently and in fashion blinded to each other’s calls and any other information regarding the specimens or strains used. Banding calls were assessed between readers, as well as resultant differences in final diagnostic calls. Excluding the CC- and AC-bands, and including the TUB-band, gene locus control-bands and gene-specific wildtype- and mutant-bands, there are 25 possible bands per MTBDR*sl* strip. There are hence 450 possible bands total for the 18 samples in the dilution series and 1300 possible bands for the 52 clinical isolates. Each strip results in two diagnostic calls and there are hence 36 possible diagnostic calls in total for 18 samples in the dilution series and 104 possible diagnostic calls in total for the 52 clinical isolates.

### Follow-up survey of TB diagnostic and research laboratories

We re-surveyed prior respondents (n=29) to our initial MTBDR*plus*-focussed survey^6^ to gather information on the current MTBDR*sl* conditions. We also surveyed for the first-time other laboratories newly-known to us to do MTBDR*sl* on smear-negative specimens (n=11). Initial non-responders were re-contacted at least twice. Survey questions included whether ramp rate changed and impact on non-valid results (survey in **Supplementary Material**). Permission to use data in an anonymised manner was obtained.

### Statistical analyses

Analyses was done using Stata version 15 (StataCorp) and GraphPad Prism version 8.0.1 (GraphPad Software) using 2-sided tests with α=0.05. McNemar’s test was used to calculate differences for paired data (i.e., the same DNA tested at both ramp rates). The two-sample proportion test was used for comparisons between proportions.

### Data availability

Available from the corresponding author.

## Results

### *MTBDR*sl *on the dilution series at different ramp rates*

Overall, irrespective of ramp rate, MTBDR*sl* did not classify the XDR-TB strain correctly at 10^2^ CFU/ml across all replicates. There were overall no differences between ramp rates of 2.2°C/s and 4.0°C/s for TUB-band detection [16/18 (89%) vs. 17/18 (94%) p=0.547], indeterminate results [2/16 (13%) vs. 3/17 (18%); p=0.680], incorrect banding calls [22/400 (6%) vs. 33/425 (8%); p=0.193)], or incorrect drug resistance calls [2/32 (6%) vs. 2/34 (6%); p=0.950] (**Table 1**). Therefore, valid results did not differ significantly [14/16 (88%) vs. 14/17 (82%); p=0.680].

**Table 1.** MTBDR*sl* performance on a dilution series of drug susceptible- and XDR-TB strains (10^4^, 10^3^, 10^2^ CFU/ml) at ramp rates of 2.2°C/s (manufacturer-recommended) or 4.0°C/s (3 replicates in triplicate for each ramp rate − 18 total MTBDR*sl* results). Accuracy for TUB-band DNA and then further analysis of indeterminate rates, incorrect banding calls and incorrect diagnostic calls were done. No significant differences were seen between ramp rates using dilution series. P-values are for within-column comparisons between different ramp rates. Data are n/N (%).

| Ramp rate<br>(°C/s) | TUB-band<br>positive | TUB-band positives |  |  |  |
| --- | --- | --- | --- | --- | --- |
|  |  | Indeterminate | Incorrect<br>banding call | Incorrect<br>diagnostic call | Valid result |
| 2.2 | 16/18*<br>(89) | 2/16<sup>†</sup><br>(13) | 22/400<sup>‡</sup><br>(6) | 2/32<br>(6) | 14/16<sup>†</sup><br>(88) |
| 4.0 | 17/18*<br>(94)<br>p=0.547 | 3/17<sup>†</sup><br>(18)<br>p=0.680 | 33/425<sup>§</sup><br>(8)<br>p=0.193 | 2/34<sup>¶</sup><br>(6)<br>p=0.950 | 14/17<sup>†</sup><br>(82)<br>p=0.680 |

\*2 strains × 3 replicates × 3 dilutions
<sup>†</sup>TUB-positive strips
<sup>‡</sup>16 TUB-band positive strips × 25 bands per strip
<sup>§</sup>17 TUB-band positive strips × 25 bands per strip
<sup>||</sup>16 TUB-band positive strips × 2 drug class diagnostic calls
<sup>¶</sup>17 TUB-band positive strips × 2 diagnostic calls
Definitions: TUB-band positive □ positive for *Mycobacterium tuberculosis*-complex DNA; indeterminate □ one or more gene locus control is absent; incorrect banding call □ the presence or absence of a band deviating from the true banding call; incorrect diagnostic call □ the presence or absence of banding patterns resulting in deviation of the true susceptibility to a drug class; valid result □ TUB-band positive, determinate for all gene locus controls, thus having diagnostic calls for both drug classes.

### *MTBDR*sl *on clinical sputa at different ramp rates*

No TUB-band detection differences were seen at 2.2°C/s vs. 4.0 °C/s [52/52 (100%) vs. 51/52 (98%), p=0.315; one MDR-TB patient was TUB-negative only at 4.0°C/s], however, indeterminate rates improved at 2.2°C/s [0/52 (0%) vs. 7/51 (14%); p=0.006] as did the proportion of bands that appeared incorrectly [0/1300 (0%) vs. 55/1275 (4%); p<0.001)] and drug-resistance calls [0/104 (0%) vs. 6/102 (6%); p=0.013] (**Table 2**). The proportion of patients with a valid result was hence 52/52 (100%) vs. 41/51 (80%). In other words, the patients who successfully received testing for FQs and SLIDs thus improved +21% (95% CI 8-34%, p<0.001).

**Table 2.**
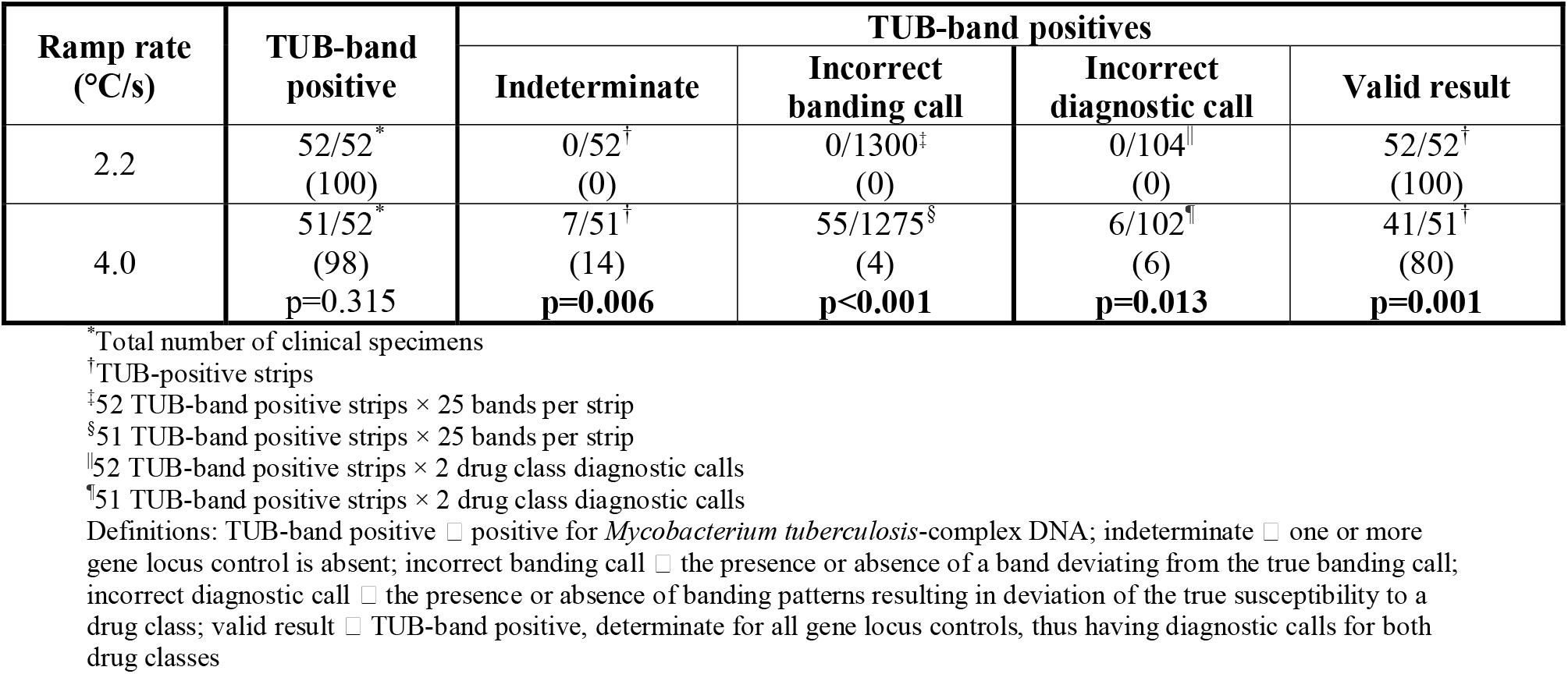
MTBDR*sl* performance on smear-negative sputa at ramp rates of 2.2°C/s (manufacturer-recommended) or 4.0°C/s (52 isolates). Accuracy for *Mycobacterium tuberculosis*-complex DNA (TUB-band), and then further analysis of indeterminate rates, incorrect banding calls and incorrect diagnostic calls were done. The number of valid results [52/52 (100%) vs. 41/51 (80%)] improved by 21% (95% CI: 8-34%; p<0.001). P-values are for within-column comparisons between different ramp rates. Data are n/N (%).

Programmatic Ultra semi-quantitative data was available for 41/52 (79%) of sputa. When load in sputa that gave a valid result at 2.2°C/s was compared to samples that gave a valid result at 4.0°C/s, there were no significant differences [median (IQR) C_Tmin_ 18.7 (17.7-19.9) vs. 18.8 (18.0-19.9); p=0.899]. We had expected 2.2°C/s to result in an improved limit of detection in MTBDR*sl* (better ability to detect higher C_Tmin_ sptua), however, we were not able to detect differences.

### Laboratory savings

If we apply the improvement in FQ and SLID testing due to optimal ramp rate usage, there would be a 21% decrease in the number of tests required to be done indirectly (which would require culture and a second MTBDR*sl*). At our local reference laboratory, ∼320 MTBDR*sl*s initially attempted on smear-negative sputum are done per month are subsequently repeated on culture isolates. Hence, in a scenario where this laboratory was using an incorrect ramp rate and changed to the correct rate, they would do ∼67 fewer indirect MTBDR*sl* tests per month would need to be repeated. At a total per test cost of USD 60 (6% per annum inflation)^12^ this translates to a saving of $48 240 per year (only factoring in pure laboratory costs).

### Inter-reader agreement

In the dilution series, diagnostic calls did not differ between the three readers at either ramp rate – all readers incorrectly classified the XDR-TB strain (as either TUB-band negative or indeterminate) at all 10^2^CFU/ml replicates and the DS-TB strain (as indeterminate) at one of the three replicates at 10^2^CFU/ml (**Table 3**). The proportion of disagreement between readers (banding calls) did not differ at suboptimal vs. optimal ramp rates [for the DS (1/225 vs. 0/225; p=0.317) or XDR strain (3/225 vs. 1/225; p=0.313)].

**Table 3.**
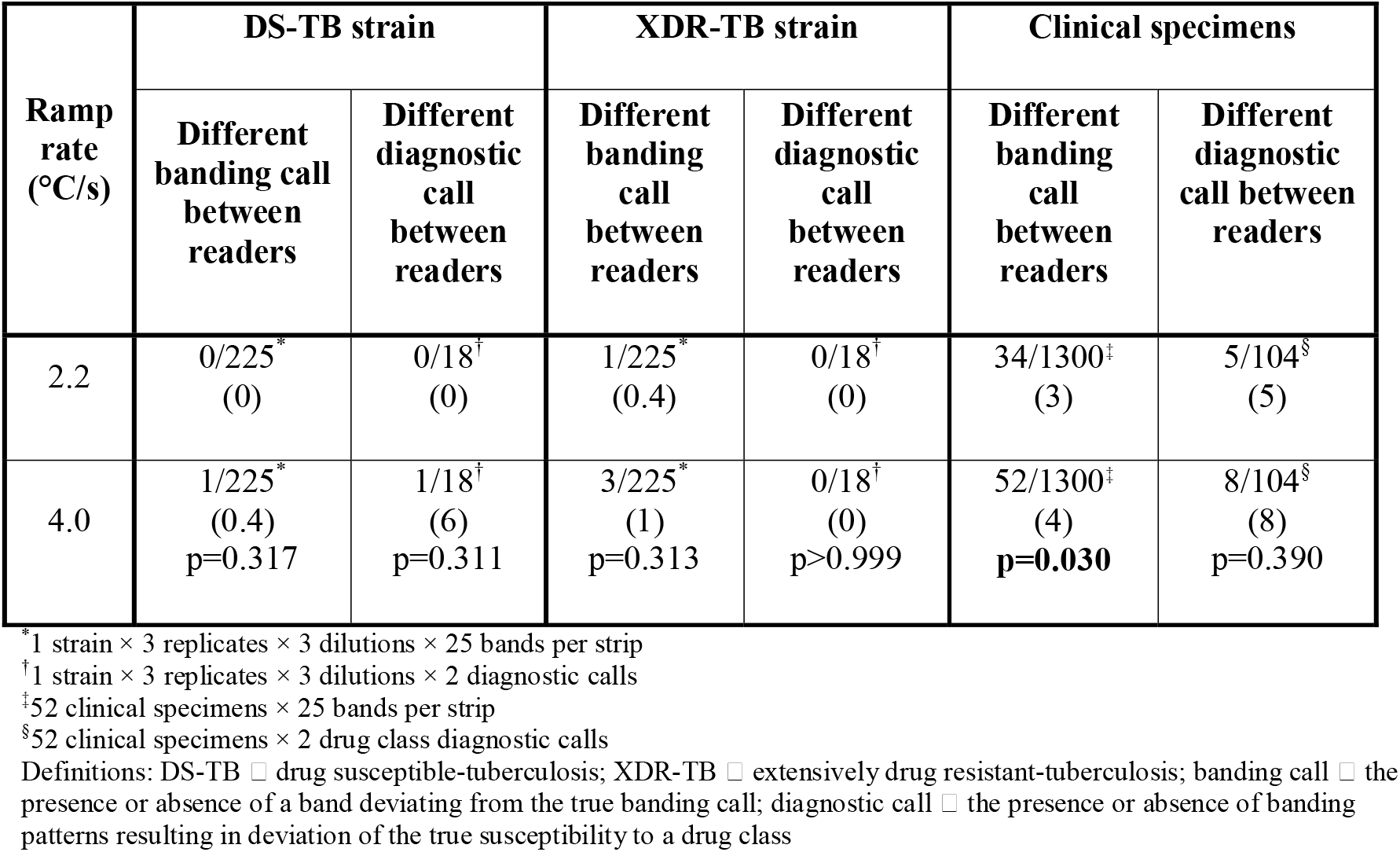
Comparison of banding and drug susceptibility calls done on a dilution series of drug susceptible- and XDR-TB strains and clinical specimens interpreted by three experienced readers. Differences in banding calls or diagnostic calls did not differ between the three readers at either ramp rate for the dilution series of cells, neither did the drug susceptibility calls in the clinical specimens, however, significant difference between readers for banding calls on the clinical sputa occurred. P-values are for within-column comparisons between different ramp rates. Data are n/N (%).

| Ramp rate (°C/s) | DS-TB strain |  | XDR-TB strain |  | Clinical specimens |  |
| --- | --- | --- | --- | --- | --- | --- |
|  | Different banding call between readers | Different diagnostic call between readers | Different banding call between readers | Different diagnostic call between readers | Different banding call between readers | Different diagnostic call between readers |
| 2.2 | 0/225*<br>(0) | 0/18†<br>(0) | 1/225*<br>(0.4) | 0/18†<br>(0) | 34/1300‡<br>(3) | 5/104§<br>(5) |
| 4.0 | 1/225*<br>(0.4)<br>p=0.317 | 1/18†<br>(6)<br>p=0.311 | 3/225*<br>(1)<br>p=0.313 | 0/18†<br>(0)<br>p>0.999 | 52/1300‡<br>(4)<br>**p=0.030** | 8/104§<br>(8)<br>p=0.390 |

\*1 strain × 3 replicates × 3 dilutions × 25 bands per strip
†1 strain × 3 replicates × 3 dilutions × 2 diagnostic calls
‡52 clinical specimens × 25 bands per strip
§52 clinical specimens × 2 drug class diagnostic calls
Definitions: DS-TB □ drug susceptible-tuberculosis; XDR-TB □ extensively drug resistant-tuberculosis; banding call □ the presence or absence of a band deviating from the true banding call; diagnostic call □ the presence or absence of banding patterns resulting in deviation of the true susceptibility to a drug class

In clinical sputa, however, although the disagreement in diagnostic calls did not differ between readers at the optimal vs. suboptimal ramp rate [5/104 (5%) vs. 8/104 (8%); p=0.390] banding calls, however, did [34/1300 (3%) vs. 52/1300 (4%); p=0.030].

### Additional survey

Twenty-nine follow-up surveys were sent to the original respondents and 11 to new laboratories. 13 total responses were received (45%), including four from new respondents (**Figure 2)**. 2/13 (15%) of respondents already had their ramp rate at 2.2°C/s (per their response to our first survey) and 6/13 (46%) had subsequently changed their ramp rate to 2.2°C/s after we communicated our previous findings^6^. Concerningly, 5/13 (39%) had not changed, for which varied reasons were offered (**Table 4**). Of the laboratories who changed to 2.2°C/s, 4/6 (67%) reported that this resulted in an improvement in banding intensity and fewer non-valid results for MTBDR*plus* and MTBDR*sl*.

**Table 4.** Laboratories that indicated their ramp rate had not yet changed to the manufacturer-recommended ramp rate of ≤2.2°C/s since the last survey, the reason why, and total number of line probe assays done per month. These laboratories do either MTBDR*plus*, MTBDR*sl* or both on smear-negative specimens but data on the subtotals for each assay were not collected.

| Country | Reason given | Number of line probe assays done per month by this respondent laboratory |
| --- | --- | --- |
| Kenya | Do not know | 240 |
| South Africa | Ramp rate change was not necessary as MTBDR*plus* assays are done on cultured isolates only and no MTBDR*sl* assays are done, as well as any changes to a standard operating procedure requires a validation process | 40 |
| Belarus | Ramp rate change in a standard operation procedure is not permitted without a prior approval process | 155 |
| Denmark | Ramp rate was not changed due to the run time of the original amplification protocol being faster | 25 |
| Spain | The thermocycler did not permit a ramp rate change | 12 |

**Figure 2.**
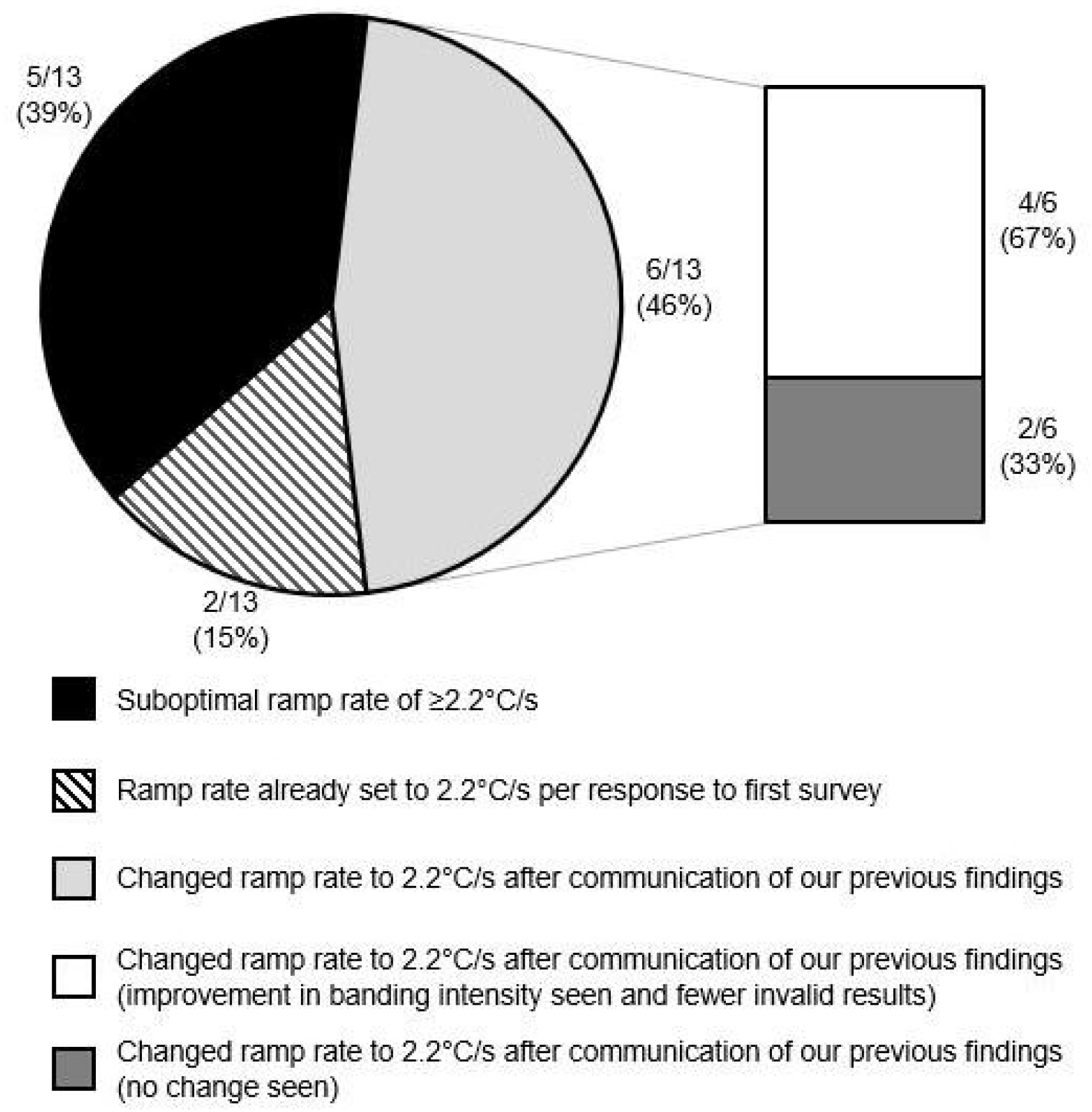
Follow-up survey results summarising thermocycler ramp rates for MTBDR*sl*. 2/13 (15%) of initially surveyed laboratories already had their ramp rate set to 2.2°C/s and 5/13 (39%) were still using a suboptimal ramp rate of ≥2.2°C/s upon resurveying. 6/13 (46%) of laboratories had, since our first survey on MTBDR*plus*, changed MTBDR*sl* ramp rate to the recommended ramp rate. Of these, 4/6 (67%) reported an improvement in banding intensity and fewer invalid results.

## Discussion

We evaluated for first-time thermocycler ramp rate’s impact on the most widely used molecular test for second-line drug-resistant TB (MTBDR*sl*). We show: 1) in sputa valid results improved by 21% when using the optimal ramp rate, which results in significant laboratory cost savings and would decrease diagnostic delay, 2) banding call and drug susceptibility call reader disagreement worsened at the suboptimal ramp rate, and 3) several laboratory respondents had not corrected their LPA ramp rate but those that had reported fewer non-valid results from MTBDR*sl* on smear-negative specimens.

We had previously shown suboptimal thermocycler ramp rate negatively affects the diagnostic accuracy of potentially thousands of MTBDR*plus* assays, especially on smear-negative sputa^6^, and ramp rate monitoring was incorporated in laboratory quality control and training documentation^7^. Now we show that a 21% increase of MTBDR*sl* diagnoses (valid results) in smear-negative specimens is possible through ramp rate correction. This is not a niche problem – we identified diagnostic laboratories who still do not do MTBDR*sl* correctly. This correction, which we have now provided MTBDR*sl*-specific empirical evidence could reduce DR-TB diagnostic care cascade gaps: a recent study found that only 65% of MDR-TB cases were evaluated for FQ resistance^13^.

Critically, ramp rate correction will reduce repeat MTBDR*sl* testing on isolates. Most directly, this will translate into substantial laboratory cost savings in high burden countries, especially when TB services are fragile due to the COVID-19 pandemic, not to mention the myriad of other individual- and population-benefits that can stem from improved DST^14^, including reduced time to treatment, transmission, and mortality.

Most laboratories in our follow-up survey had corrected ramp rate, however, a significant amount, including those responsible for routine diagnostic testing on smear-negative specimens, still used a suboptimal ramp rate. We re-iterate that 1) laboratories ensure that they are using the optimal ramp rate, 2) thermocycler ramp rate monitoring be added to laboratory external quality assurance programmes and accreditation processes for MTBDR*sl*, and 3) that the manufacturer makes the recommended ramp rate more prominent in assay documentation. It is worth evaluating further why incorrect ramp rates continued to be used. This may be due to quality assurance lapses, a deliberate choice (e.g., to potentially speed up turn-around-time) without an awareness of downsides, or a design limitation of available thermocyclers.

We saw a more prominent performance difference between ramp rates in clinical sputa than in spiked solution. Bacilli in mucus sputa matrices behave differently to bacilli spiked in *in vitro* experiments and our findings illustrates potential downsides to investigating the effect of PCR parameters on molecular assays when *in vitro* or mock specimens are used.

Our evaluation has strengths and limitations. We did not assess a wider ramp rate range due to limited sputa and cost but used the most frequently reported incorrect ramp rate. Our survey results would have also been subjected to selection, response and reporting biases and we suggest a formal survey is done by the manufacturer and/or the appropriate regulatory and oversight agency (we did our survey independently). We did not evaluate savings stemming from quicker diagnosis, treatment initiation, and long-term reductions in transmission and mortality due to improved performance – there is already a saving in laboratory costs alone with no downside.

In conclusion, we have shown that this incorrect and seemingly innocuous technical setting (ramp rate) has a real-world negative impact on patients’ diagnoses for second-line drug resistance using MTBDR*sl*. Smear-negative patients are especially vulnerable. All stakeholders must ensure that the optimal thermocycler ramp rate for MTBDR*sl* is used, and this requires investigation for other molecular diagnostics.

## Data Availability

Available from the corresponding author.

## Acknowledgments

The authors thank the National Health Laboratory Services, Green Point, Cape Town, South Africa, and Hain Lifescience, Germany.

The authors thank the laboratories that participated in the survey and provided data.

## Author Contributions

B.D., M.dV., G.T. and R.W. conceived the experiments. T.D. and S.P. provided specimens and data from the National Health Laboratory Service, Greenpoint. B.D. conducted the experiments and analysed the data. S.P, Y.G., R.V. and S.M. assisted with analysis of results. J.M. provided critical input. All authors reviewed the manuscript.

