## Supplementary Material for "Frequent suboptimal thermocycler ramp rate usage negatively impacts MTBDR*sl* performance for second-line drug resistant tuberculosis diagnosis"

**Supplement**

**MTBDR*sl* thermocycler ramp rate survey sent to laboratory respondents who completed our first survey on MTBDR*plus^1^***

This new survey was also sent to laboratories known to be doing line probe assays on smear‑negative specimens but not originally contacted.

1. Name
2. Name of your organisation
3. Email address
4. What country is your laboratory based in?
5. Since completing the previous survey, did your laboratory change the thermocycler ramp rate to 2.2°C/s when doing MTBDR*plus* and/or MTBDR*sl*?
   - Yes
   - No
6. When did your laboratory change the ramp rate? (mm/yyyy format)
7. If the answer was no to question 5, why did your laboratory NOT change the ramp rate to 2.2°C/s? (Please complete question 8 and 9 then skip to the end of the survey and click submit)
   - Thermocycler did not permit ramp rate change
   - Do not know
   - Changes to test SOP are not permitted without a prior approval process
   - Thermocycler ramp rate was already set to 2.2°C/s
   - Other:_____________________________________________________________
8. Did your laboratory change any other MTBDR*plus* and/or MTBDR*sl* test parameters?
   - Yes
   - No
9. If yes to question 8, please elaborate:
10. Did the correction of ramp rate result in an improvement in banding intensity when doing MTBDR*plus* and/or MTBDR*sl*?
    - Yes
    - No
11. If yes to question 10, please elaborate (e.g. are there specific bands that have improved the most (including control bands):
12. Did the correction of ramp rate result in fewer non-actionable (TUB band-negative or indeterminate for any drug locus control band) MTBDR*plus* results?
    - Yes
    - No
    - Do not know
    - Other:_____________________________________________________________
13. Did the correction of ramp rate result in fewer non-actionable (TUB band-negative or indeterminate for any drug locus control band) MTBDR*sl* results?
    - Yes
    - No
    - Do not know
    - Other:_____________________________________________________________
14. In the 3 months BEFORE your laboratory corrected the ramp rate, is there data available on how many MTBDR*plus* tests were done directly on smear-negative sputum?
    - Yes
    - No
      i. If no to question 14, why?
         - Not recorded
         - Too difficult to retrieve data
         - Other:_________________________________________________
      ii. If yes to question 14, how many MTBDR*plus* tests were done directly on smear-negative sputum?
      iii. How many of these, done directly on smear-negative sputum, were TUB-band negative (TB-negative)?
      iv. How many of these, done directly on smear-negative sputum, were TUB-band positive, but indeterminate for any gene locus (*rpoB*, *katG* or *inhA*)?
15. In the 3 months BEFORE your laboratory corrected the ramp rate, is there data available on how many MTBDR*sl* tests were done directly on smear-negative sputum?
    - Yes
    - No
      i. If no to question 15, why?
         - Not recorded
         - Too difficult to retrieve data
         - Other:­­­­­­­­­­­­­­­­­­­­­­_________________________________________________
      ii. If yes to question 15, how many MTBDR*sl* tests were done directly on smear-negative sputum?
      iii. How many of these, done directly on smear-negative sputum, were TUB-band negative (TB-negative)?
      iv. How many of these, done directly on smear-negative sputum, were TUB-band positive, but indeterminate for any gene locus (*gyrA*, *gyrB*, *rrs* or *eis*)?
16. In the 3 months AFTER your laboratory corrected the ramp rate, is there data available on how many MTBDR*plus* tests were done directly on smear-negative sputum?
    - Yes
    - No
      i. If no to question 16, why?
         - Not recorded
         - Too difficult to retrieve data
         - Other:­_________________________________________________
      ii. If yes to question 16, how many MTBDR*plus* tests when done directly on smear-negative sputum?
      iii. How many of these, done directly on smear-negative sputum, were TUB-band negative (TB-negative)?
      iv. How many of these, done directly on smear-negative sputum, were TUB-band positive, but indeterminate for any gene locus (*rpoB*, *katG* or *inhA*)?
17. In the 3 months AFTER your laboratory corrected the ramp rate, is there data available on how many MTBDR*sl* tests were done directly on smear-negative sputum?
    - Yes
    - No
      i. If no to question 17, why?
         - Not recorded
         - Too difficult to retrieve data
         - Other:_________________________________________________
      ii. If yes to question 17, how many MTBDR*sl* tests were done directly on smear-negative sputum?
      iii. How many of these, done directly on smear-negative sputum, were TUB-band negative (TB-negative)?
      iv. How many of these, done directly on smear-negative sputum, were TUB-band positive, but indeterminate for any gene locus (*gyrA*, *gyrB*, *rrs* or *eis*)?
